# The role of Zambia’s expansive Inter-agency Coordinating Committee (ICC) in supporting evidence-based vaccine and health sector programming

**DOI:** 10.1101/2021.11.29.21267021

**Authors:** Zoe Sakas, Katie Rodriguez, Kyra A Hester, Roopa Darwar, Bonheur Dounebaine, Anna S Ellis, Simone Rosenblum, Kimberley R Isett, Walter Orenstein, Matthew C Freeman, William Kilembe, Robert A Bednarczyk

## Abstract

New vaccines, technologies, and regulations, alongside increased demand for vaccines, all require prioritization and coordination from key players within the vaccine sector. Inter-agency Coordinating Committees (ICC) support decision-making and coordination at the national-level and act as key drivers for sustainable improvements in vaccination programming. We employed a qualitative case study design to investigate critical success factors for the high routine immunization coverage in Zambia from 2000-2018. Qualitative data were collected between October 2019 and February 2020, including key informant interviews (n=66) at the national, provincial, district, and health facility levels. Thematic analysis was applied to understand the role of the Zambian ICC and its impact on the policy environment overtime. Throughout our study period, the Zambian ICC demonstrated the following improvements: 1) expanded membership to include diverse representation; 2) expanded scope and mandate to include maternal and child health in decision-making; and 3) collaboration with the ZITAG through distinct roles. The diverse and expansive membership of the Zambian ICC, along with its ability to foster government commitment and lobby for additional resources, supported improvements in immunization programming. The Zambian ICC holds considerable influence on government agencies and external partners which facilitates procurement of funding, policy decisions, and strategic planning.

## 1. Introduction

Immunization policies and programs in low and middle-income countries are constantly evolving.[1] The introduction of new vaccines, technologies, and regulations, alongside increased demand for vaccines, all require prioritization and coordination from key players within the vaccine sector.[1] To respond to growing needs, the establishment and maintenance of effective, efficient, and rigorous national-level coordination forums are essential.

In the 1990s, the World Health Organization (WHO) established Inter-Agency Coordinating Committees (ICCs) as country-level forums to coordinate polio eradication programming.[2] In 2006, Gavi, the Vaccine Alliance (Gavi) began using ICCs to support broader management of funds for immunization programming. [3,4] Gavi also requires creation of National Immunization Technical Advisory Groups (NITAG) to present data and evidence-based recommendations to the ICC and other national forums. [5,6,7] Since 2001, Gavi has contributed $164,000,000 to the Zambian government for vaccination programming and health systems strengthening, which has supported the expansion and strengthening of Zambia’s ICC and NITAG, referred to as ZITAG. [7, 8]

Literature suggests that ICCs and NITAGs should have distinct roles – the ICC supports supervision and coordination, and the NITAG provides technical expertise to promote rigorous decision-making [1,7]. However, relying on parallel administrative structures may foster confusion around roles and responsibilities, leading to inefficiencies and disruptions in program functioning [6,9,10]. Tailoring the roles and responsibilities of these unique forums to ensure they complement, rather than disrupt, each other may help overcome these challenges.

Successful coordination forums act as key drivers for sustainable improvements in vaccination programming and coverage [1,5,11]. This study illustrates how Zambia bolstered its ICC for long-term functionality, expanded ICC membership and scope beyond Gavi requirements, and distinguished a complementary structure for the Zambian Immunization Technical Advisory Group (ZITAG). In Zambia, stakeholders reported that this approach worked well to support immunization programming and improve vaccination coverage [11].

## 2. Methods

This study is nested within a larger research project which investigates key success factors for improvements in routine immunization coverage in Zambia, Nepal, and Senegal.

**Research questions:**

1. What were key drivers of success for high and sustained routine immunization coverage?

1. What were the key implementation and change management strategies employed for high growth in vaccine coverage levels?
2. How has the policy and decision-making environment changed over time, and how have these changes (or lack thereof) impacted both the functioning of the vaccination program and the environment in which it works?

1. How are country wide policy decisions made?
2. What is the interplay of the NITAG and ICC, and how does this top-down leadership impact decision-making?

This paper details one of the most influential success factors described by key informants – the role of the Zambian ICC – and its impact on the policy environment overtime.

### 2.1 Study design and setting

We employed a qualitative case study design to investigate factors that supported improvements in routine immunization coverage for children under 1 year of age in Zambia from 2000 to 2018. Zambia was one of three countries selected as exemplars in vaccine delivery [12]. Country selection for exemplars was determined by high and sustained DTP1 and DTP3 coverage (illustrated in Figures 1 and 2, respectively), which served as proxies of the vaccine delivery system. Lusaka, Central, and Luapula provinces were chosen based on vaccination coverage and contextual factors. Figure 3 illustrates routine immunization coverage improvements in different regions in Zambia to demonstrate heterogeneity between study sites [11]. Additional information about the site selection, sampling, and protocol can be found elsewhere [11,12].

**Figure 1.**
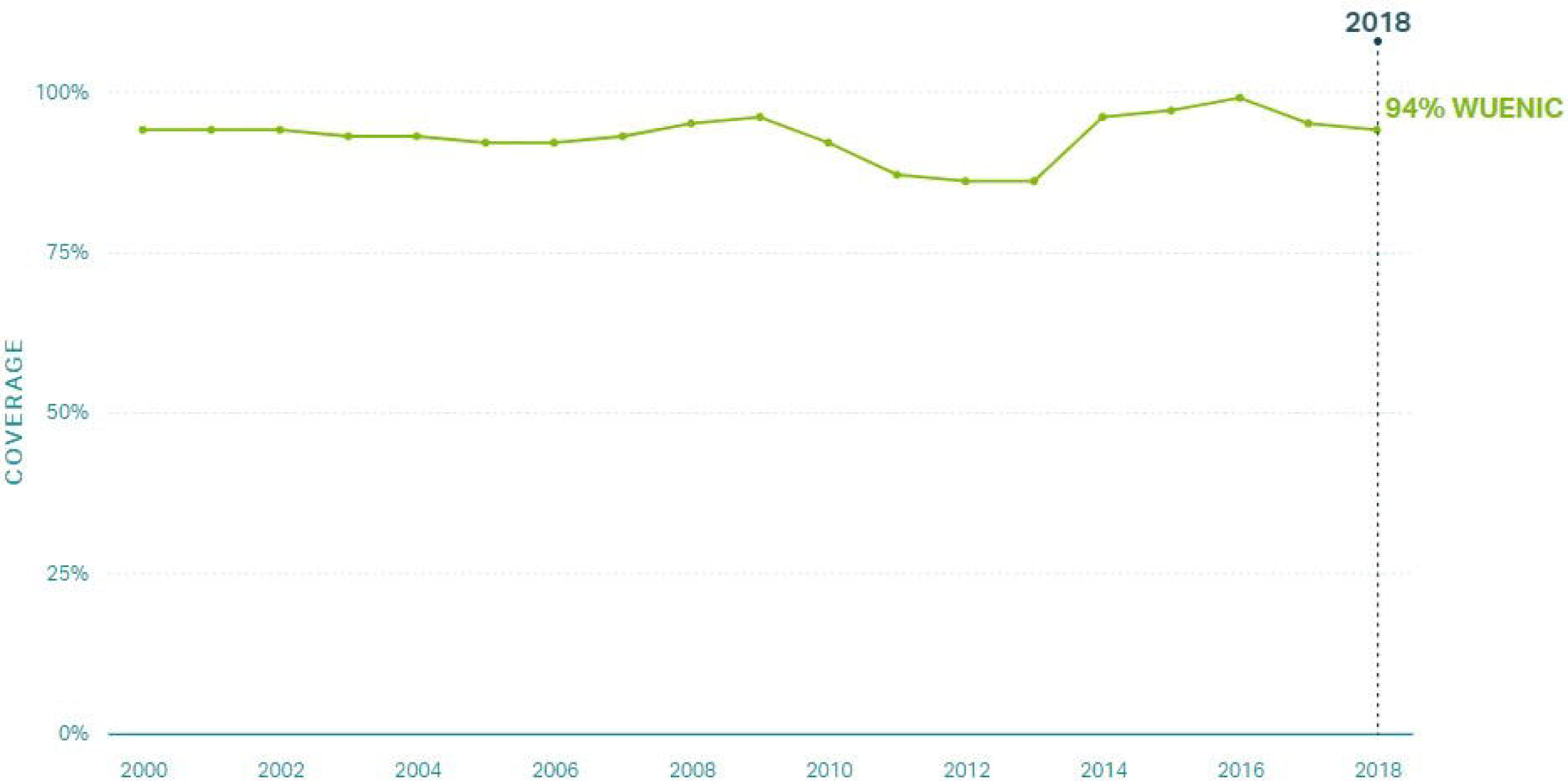
DTP1 coverage in Zambia 2000-2018.

**Figure 2.**
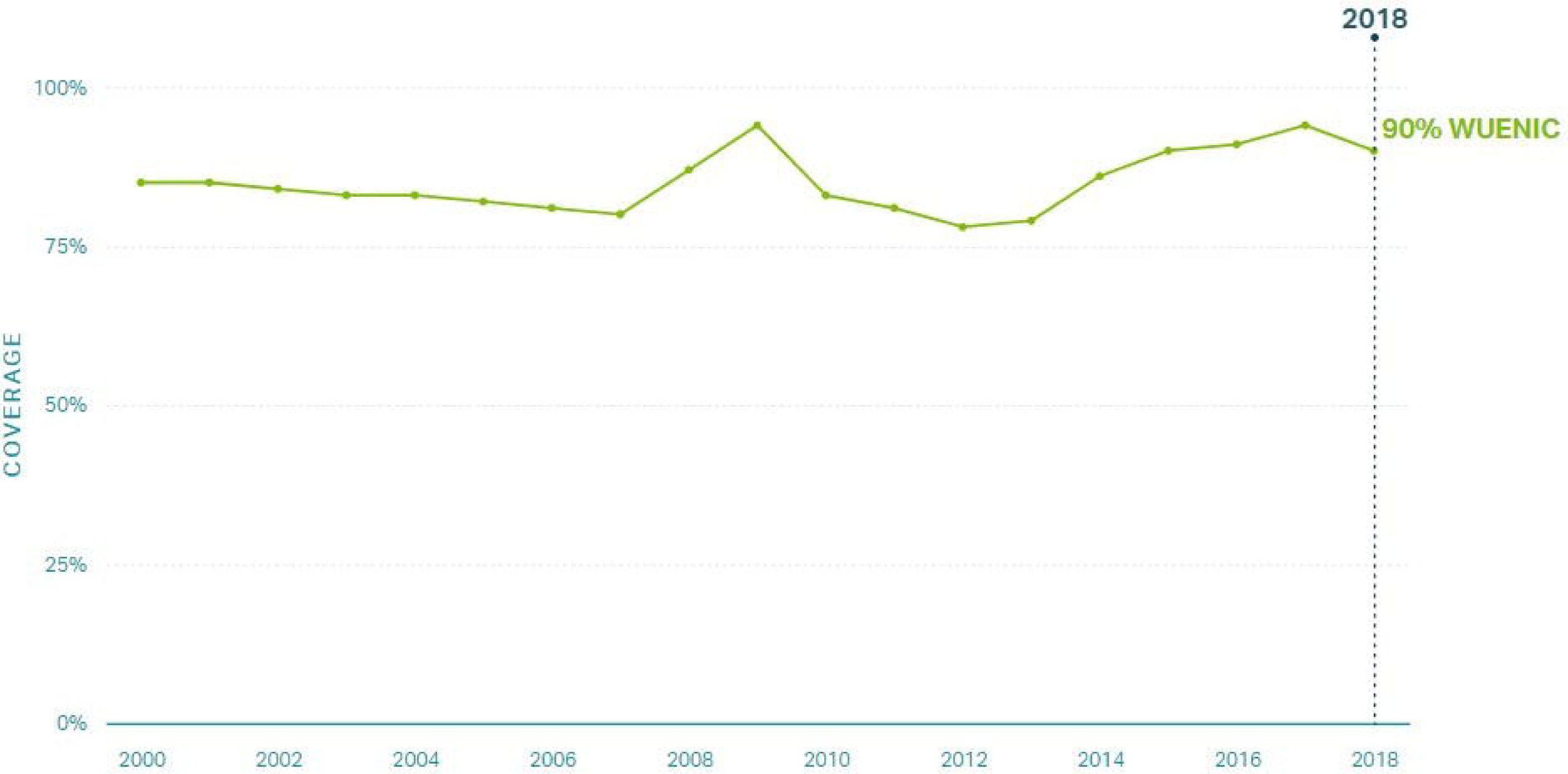
DTP3 coverage in Zambia 2000-2018.

**Figure 3.**
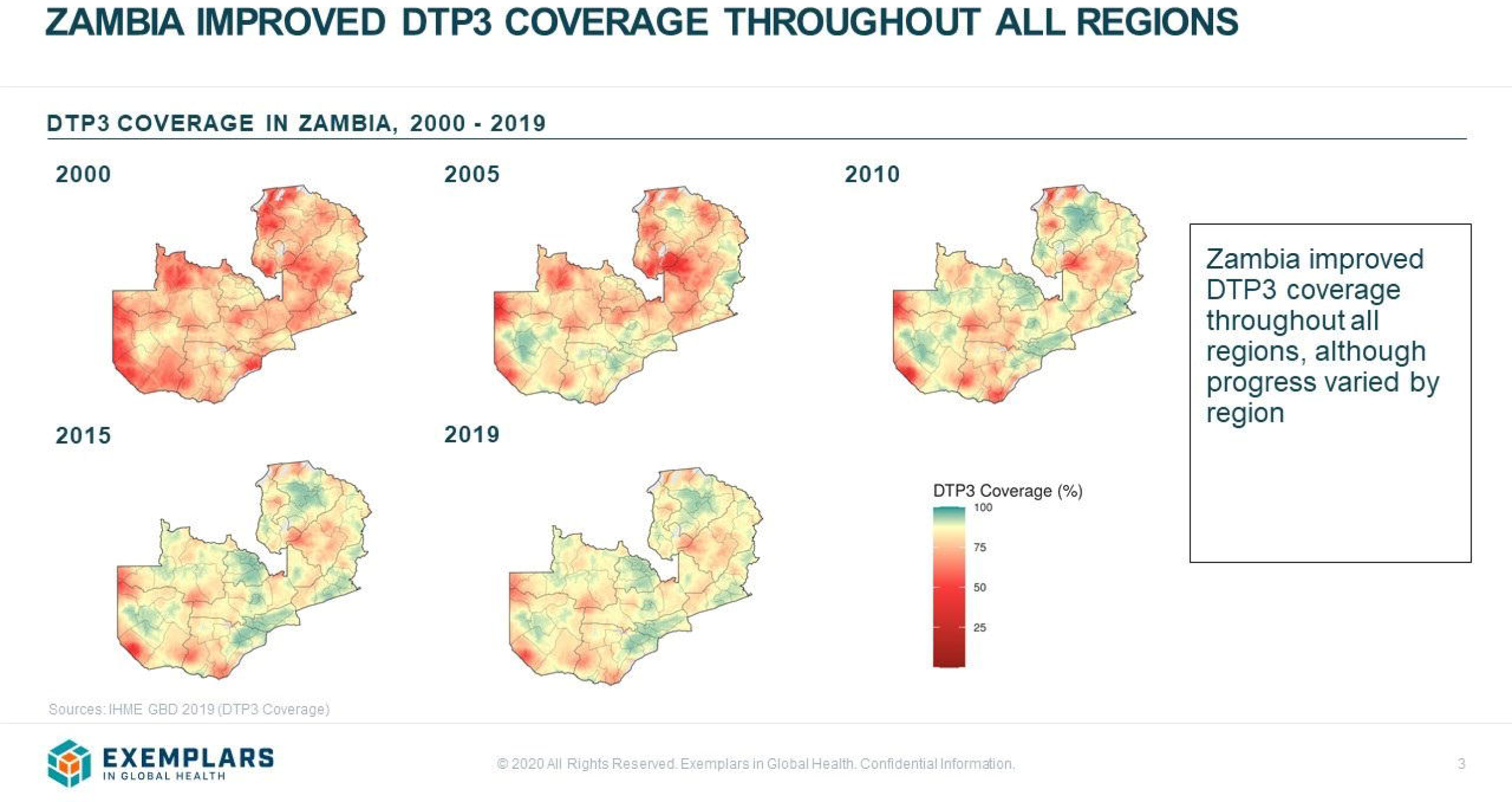
Heat map illustrating improved DTP3 coverage across all regions of Zambia 2000-2019.

### 2.2 Data collection and analysis

Qualitative data were collected between October 28, 2019 and March 6, 2020. Data collection included key informant interviews (n=66) at the national, provincial, district, and health facility levels. Data collection activities are summarized in Table 1. Data were collected by the Center for Family Health Research in Zambia (CFHRZ). Key informant interview (KII) guides were translated into Nyanja and Bemba languages by research assistants. Topics guides aimed to identify key factors that drove the success of the vaccination program in Zambia, according to key informants, especially during points of catalytic growth in DTP1 and DTP3 coverage. All interview guides were piloted before use and adjusted iteratively throughout data collection.

**Table 1:** Summary of research activities, October 2019 – February 2020.

| Method | Participants | Number of activities | Number of participants |
| --- | --- | --- | --- |
| Key Informant Interviews | Ministry of Health | 9 | 10 |
|  | Ministry of Education | 1 | 1 |
|  | Ministry of Finance | 1 | 1 |
|  | Partner organization | 11 | 15 |
|  | Provincial Health Office | 6 | 8 |
|  | District Health Offices | 10 | 19 |
|  | Nurses in-charge of health facilities | 7 | 10 |
|  | Community-based volunteers | 11 | 11 |
|  | Community leaders | 10 | 10 |
| **Total** |  | **66** | **85** |

An initial list of KIIs was developed with CFHRZ and MoH officials; we then used snowball sampling to identify additional key informants. The duration of KIIs averaged one and a half hours. KIIs were audio-recorded with the permission of participants. Research files, recordings, and transcriptions were de-identified and password protected. Debriefs were conducted during data collection to identify emerging themes and follow-up questions. All the topic guides used for this study, as well as our analysis tools, can be found on our Open Sciences Framework webpage [13].

We conducted a thematic analysis of the transcripts to identify contributing factors to the success of the immunization program. We developed a codebook using a deductive approach, applying constructs from existing implementation science frameworks, and adjusted the codebook based on the emerging themes. Transcripts were coded and analysed using MaxQDA2020 software (Berlin, Germany). All transcripts were coded, relevant themes were identified, and visual tools were used to illustrate the findings. We considered setting and participant roles while identifying key points, and further contextualized data using historical documents and a literature review. Further information about data management and analysis can be found elsewhere [11,12].

### 2.4 Gavi document review

Following qualitative data analysis, we reviewed all Gavi annual reports for Zambia, including joint appraisals, that were publicly available within our study period [5]. During this document review, we focused on themes that emerged from the qualitative data. Findings from the qualitative data were triangulated with information from Gavi documents to strengthen the analysis.

### 2.5 Research advisory group and stakeholder feedback

The findings were shared with a research advisory group composed of international and Zambian stakeholders and experts in the governance, policy, and immunization sectors. Follow-up questions were shared with in-country key informants to gain more information and detail. Details about advisory group members can be found elsewhere [12].

### 2.6 Ethics Statement

The study was approved by the University of Zambia Biomedical Research Ethics Committee (Federal Assurance No. FWA00000338, REF. No. 166-2019), the National Health Research Authority in Zambia, and the Institutional Review Board committee of Emory University, Atlanta, Georgia, USA (IRB00111474). Informed consent was obtained from all 85 participants. Written consent forms were provided, reviewed, and signed prior to interviews. All consent forms were approved by the appropriate ethics committees.

## 3. Key findings & Implications

Many key informants, especially at the national and regional levels, identified the Zambian ICC as a key success factor for increasing vaccination coverage. Additionally, the Zambian ICC was described as a *“key driver of sustainable coverage and equity”* in the 2018 Gavi joint appraisal, which strengthens the findings from our qualitative data [5]. According to our analysis, the Zambian ICC has held considerable influence on government agencies and external partners, which facilitates the procurement of funding, policy decisions, and strategic planning.

> “For the EPI [Expanded Programme on Immunization], the Inter-Agency Coordinating Committee is a very important organ for policy development and resource mobilization.” (WHO personnel)

The Zambian ICC was established in 1999 to meet WHO Polio Eradication Initiative requirements. The ICC was adapted in 2006 to support the management of Gavi funding for general immunization programming. The ICC continues to oversee vaccine initiatives, lobby for funding from the national government and partners, and foster collaboration between the Ministry of Health (MoH), partners, and community-level stakeholders [5]. The key responsibility of the ICC is to facilitate decision-making and resource mobilization through monthly meetings.

Throughout our study period, particularly from 2014 to present, the Zambian ICC has demonstrated the following improvements: 1) expanded membership to include diverse representation; 2) expanded scope and mandate to include maternal and child health in decision-making; and 3) collaboration with the ZITAG through distinct and complimentary roles and responsibilities. These factors are described further below.

### 3.1 The Zambian ICC expanded its membership and scope to improve reach, coordination, and commitment

According to key informants and Gavi joint appraisals from 2014 to 2018, the Zambian ICC performed above the minimal requirements obligatory for funding [5]. The ICC sufficiently accomplished recommendations from Gavi related to strategic planning, financing, membership, information dissemination, and decision-making procedures which are outlined in joint appraisal reports [5]. These improvements were likely fuelled by recommendations from meetings in 2016 and 2017 which included Gavi representatives, current ICC members, and key stakeholders from the Zambian government. The extent that the ICC improved from 2014 to 2018 suggests commitment and motivation from internal leadership and champions. Specifically, the Zambian ICC expanded both its membership and mandate to promote decision-making and resource allocation.

#### 3.1.1 Diverse and representative ICC membership

The Zambian ICC includes diverse and representative membership to support collaboration among technical experts, decision-makers, and community organizations [7]. With its multi-sectoral composition, the ICC provides a forum for coordination of immunization investments, bolsters management of key action points, and administers technical working groups [15,16]. The MoH holds a leadership position and is considered a “*driving force*” of the forum’s success, according to national-level stakeholders. Additionally, over 50% of Zambia’s ICC members report having been members of the ICC for over 6 years, according to a recent Gavi evaluation [7]. Diversity and longevity of ICC members was described as beneficial to decision-making.

All members of the ICC are expected to share their opinions and vote on decisions to support collaboration between entities represented on the forum [5,15,16]. Additionally, members of the ICC may participate in other forums that contribute to development in Zambia, including meetings with the MoH to develop national health strategic plans and engagement with advisory committees for other health sectors [7]. This cross-membership supports efficiency of planning and resource allocation for health and development in Zambia through the alignment of ideas and priorities between decision-making bodies. Existing literature states that members of the ICC, ZITAG, and EPI Technical Working Groups are generally informed about decisions, plans, and evaluations in Zambia [7] – which aligns with our findings.

Table 2 outlines key ICC membership required or recommended by Gavi, and compares the fulfilment of these guidelines by the Zambian ICC in 2014 and 2018 [5]. Gavi support is reliant on maintaining an active ICC, which includes at least one member in each of the required categories:

1. Ministry of Health,
2. EPI Programming,
3. Financial Planning,
4. Health Systems Strengthening,
5. Key Donors, and
6. Implementing Partners.

**Table 2:**
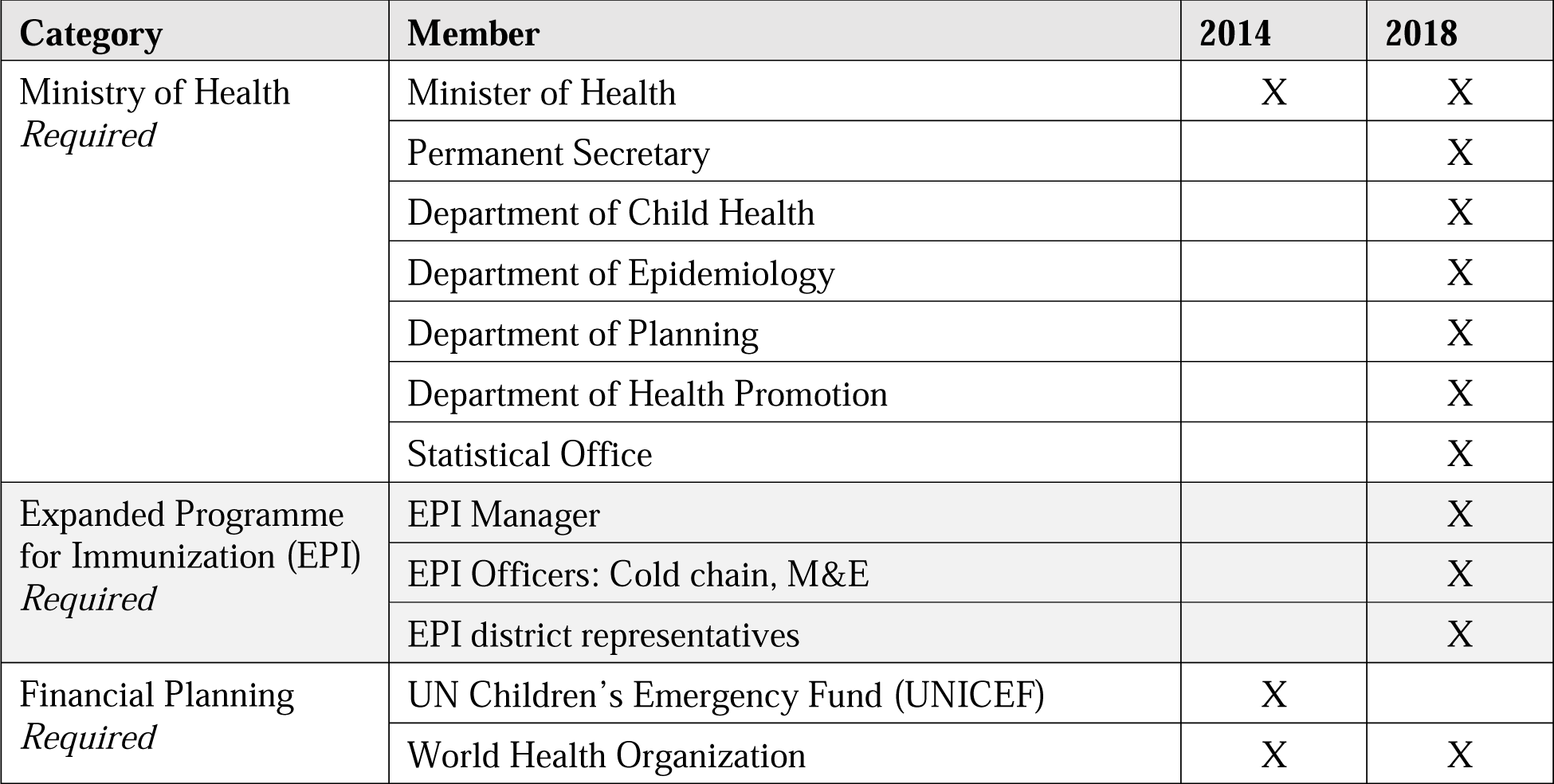
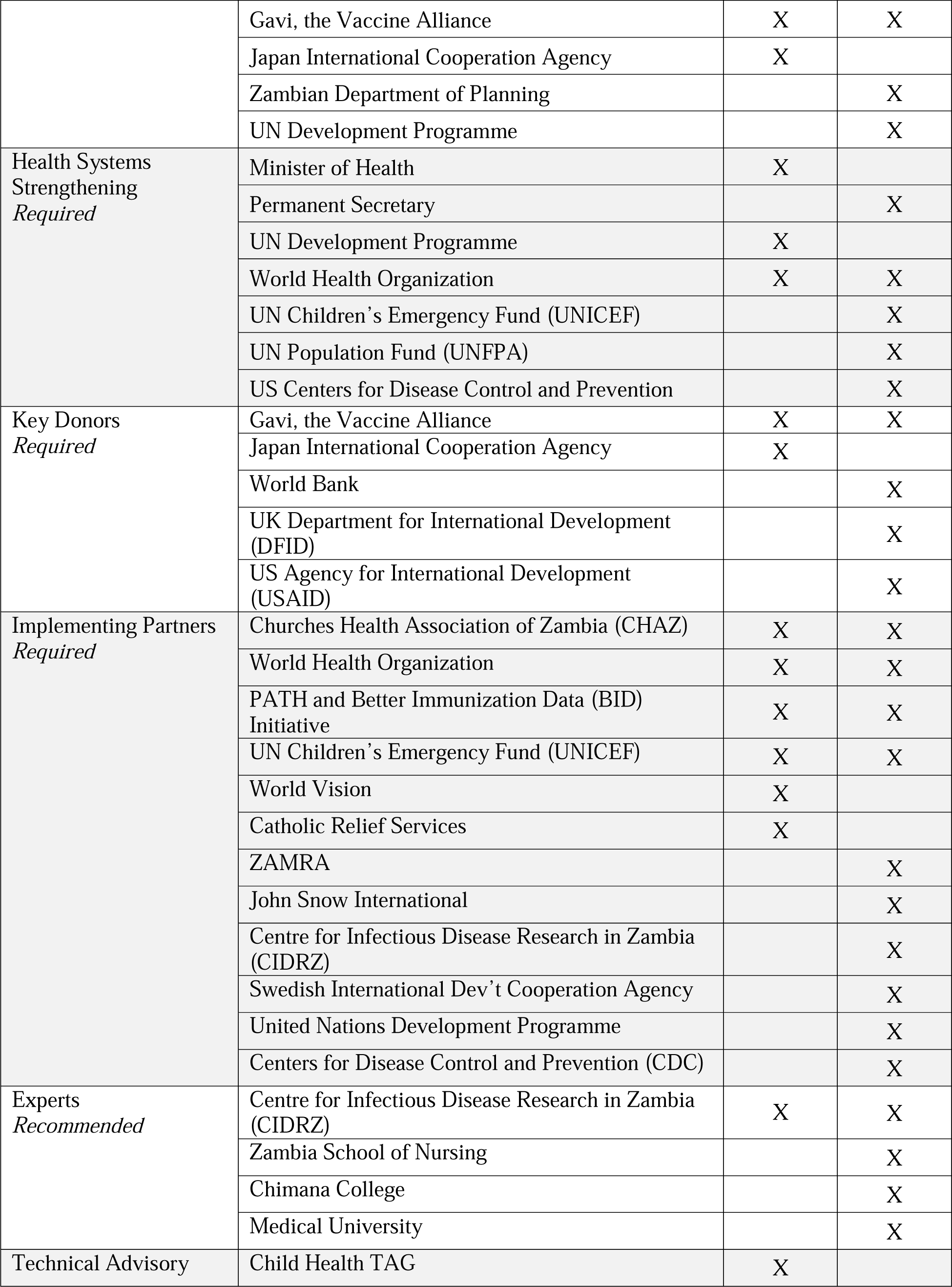
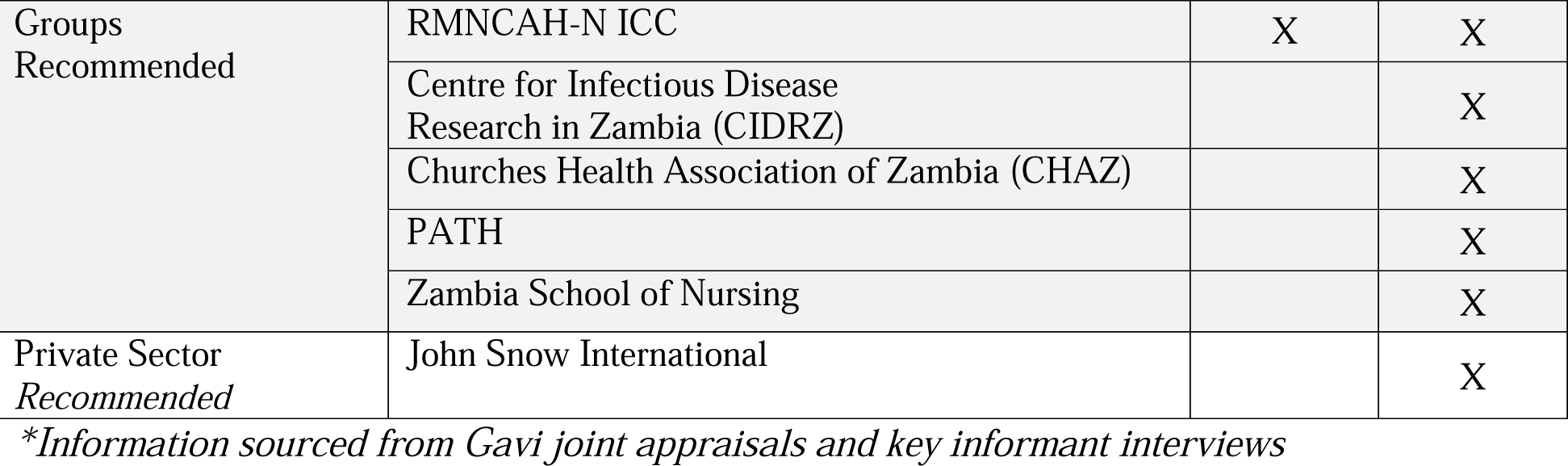
ICC membership matrix from 2014 and 2018*.

Additionally, Gavi recommendations include ICC representation from the following:

1. 7) Immunization Experts,
2. 8) Technical Advisory Groups, and
3. 9) Private Sector.

In 2018, the Zambian ICC membership filled all membership categories. The drastic change in ICC membership from 2014 to 2018, illustrated by the table below, demonstrates Zambian commitment to strengthening the ICC and improving collaboration for immunization programming. Although the membership of the ICC expanded significantly over the last decade, there are still apparent gaps, including limited private sector representation. Additionally, recent reports highlight concerns that some members were more of diplomats in their organizations and might be lacking the required technical capacity [7].

The expansive membership of the ICC affected various components of vaccination programming, including cold chain expansion and data quality, through the inclusion of representatives from sub-committees. The Expanded Program on Immunization (EPI) in Senegal includes a technical working group, which functions separately from the ICC. The EPI technical working group (EPI-TWG) is divided into four subcommittees: 1) Monitoring and evaluation, 2) Social mobilization, 3) Cold chain and logistics, and 4) Service delivery. EPI-TWG subcommittees are known to be highly active and contributed substantially to the success of the Zambian ICC, especially through providing reports and evidence to both the ICC and ZITAG. [7] A representative for each sub-committee is represented on the ICC and attends quarterly meetings for decision-making and resource mobilization. Additionally, integrated meetings, attended by ICC members and representatives from the national, provincial, and district levels, are conducted to review data and address gaps in coverage.

Membership of the ICC includes, as illustrated in Figure 4:

- External donors and implementing partners to align priorities and foster mutual trust.
- EPI officers, including representatives from all districts and sub-committees to present sector-specific updates and support context specific implementation.
- Select representatives from technical institutions, including researchers from the Centre for Infectious Disease Research in Zambia (CIDRZ), to ensure evidence is considered.

**Figure 4.**
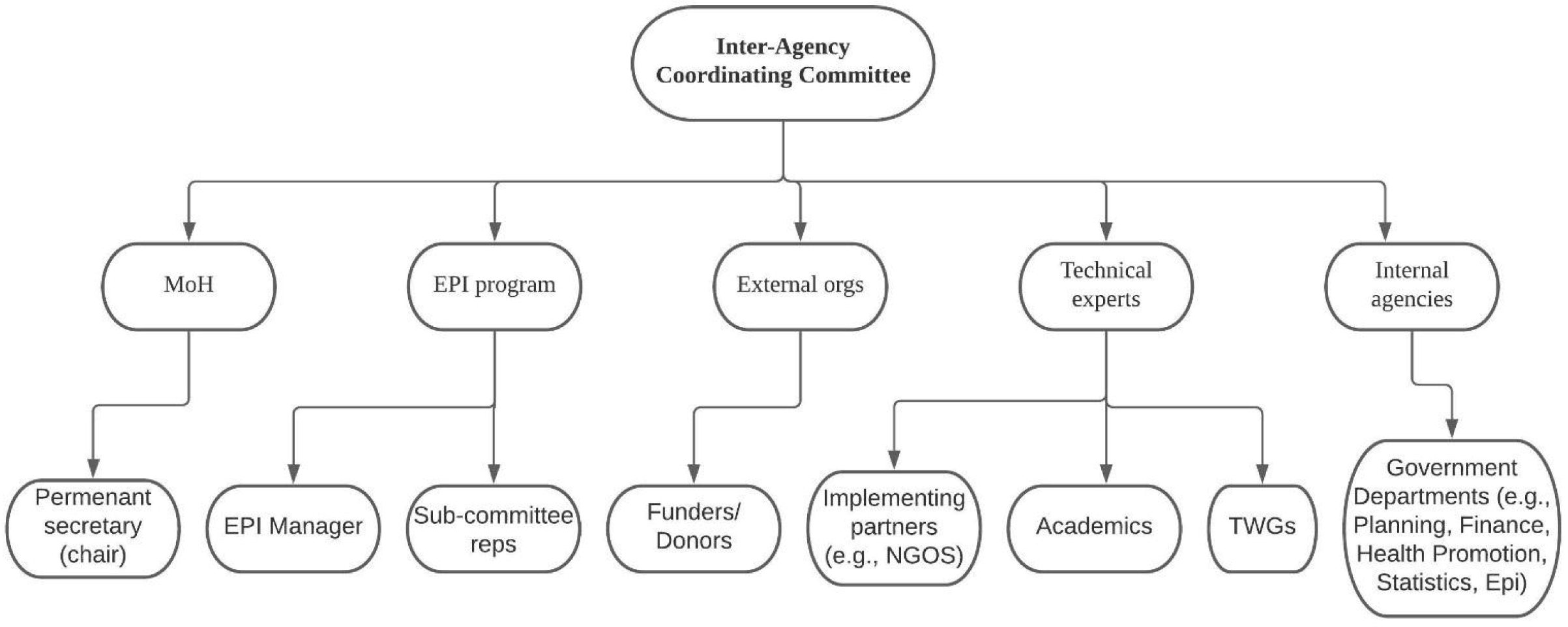
Description of the ICC structure in Zambia.

> “The ICC is used for [government officials] or partners who support programs in Zambia. It is a committee for high profile [officials], those who have interest in immunization. Together we tell them to [describe] the performance of the program and some of the challenges that it is facing, so that people can lobby for support in terms of finances and any other things that we have had tried from the program operation.” (EPI Cold Chain Officer)

#### 3.1.2 Expanded mandate to other areas of health

In 2017, the terms of reference for the Zambian ICC expanded to include a broader focus on Reproductive, Maternal, Neonatal, Child, and Adolescent Health, and Nutrition (RMNCAH-N). This decision, discussed during an annual appraisal meeting with key stakeholders, stemmed from a suggestion by Gavi to the Zambian ICC to expand its scope [5]. Following the appraisal meeting, the ICC mandate expanded to include organizations that work in child and maternal health. Key informants from the national and regional level spoke about the positive impact of this expanded mandate, which allows for efficient and comprehensive strategic planning and resource allocation.

> “In an effort to strengthen the governance and oversight capabilities of the ICC, there are recommendations to prioritize ICC strengthening in 2018 through an assessment which will include a comprehensive review of both the TORs and membership of the group.” (Gavi appraisal, 2017)

> “The ICC is ideally targeted at the heads of organizations who have an interest in reproductive and maternal and child health and nutrition issues in the country.” (UNICEF personnel)

Although Gavi appraisals stated that the Zambian ICC began to incorporate broader areas of health in 2017, there is evidence from interviews that the ICC was working to expand its scope to include maternal, new-born, and child health representation as early as 2010. This expansion allowed for a more holistic approach to vaccination, involving ante-natal clinics, traditional birth attendants, nurses, and other staff members from health facilities. Activities such as child growth tracking and school enrolment were also tied to vaccination efforts.

> “In terms of decision-making at a higher level, the ICC [has] evolved some time back. There was the RMNCAH-N partnership for maternal, newborn, and child health. It must have been 2010, there was a partnership which was created to incorporate the maternal component.” (WHO personnel)

The ICC’s motivation to include RMNCAH-N was likely in response to prioritization of women’s, children’s, and adolescent health within the global health policy environment; however, this motivation was not entirely clear from our investigation. It is possible that introduction of the Human Papillomavirus (HPV) vaccine (piloted and scaled up in Zambia in 2013 and 2019, respectively) may have supported this expansion. Representatives focused on HPV immunization were likely involved with the ICC at that time, establishing a natural connection between reproductive and vaccination programming, which highlights the importance of integrated programming and budgeting [17].

Although key informants interviewed for this study spoke positively about the expanded mandate and cross-sectoral approach, a recent Gavi evaluation revealed that members of the ZITAG and EPI-TWG had some concerns over the recent changes. [7] Namely, ZITAG members reported that the link between them and the Zambian ICC was weakened by the broadening mandate which led to inefficiencies within the immunization sector. [7] Some also described inefficiencies in decision-making related to vaccine programming due to the ICC’s broader focus. [7]

### 3.2 Distinct and complementary roles of the Zambian ICC and ZITAG

Our data illustrates that strengthening the relationship between the Zambian ICC and ZITAG for long-term functionality supported improvements in vaccine programming. However, other recent studies also highlight the challenges of coordinating between two similar national forums. [A] The ZITAG was established in 2016 to provide evidence-based recommendations and as a requirement to continue receiving Gavi funding. [7,8] To date, the ICC and ZITAG have distinct and complementary roles and responsibilities, as described in Figure 5 and Table 3. Gavi-funded programs typically focus first on the adoption of an ICC to manage donor funding, and later on the establishment of the NITAG as a parallel technical group. Key informants from the MoH described that the collaboration between the Zambian ICC and ZITAG allowed for long-term functionality without duplication of efforts.

**Figure 5.**
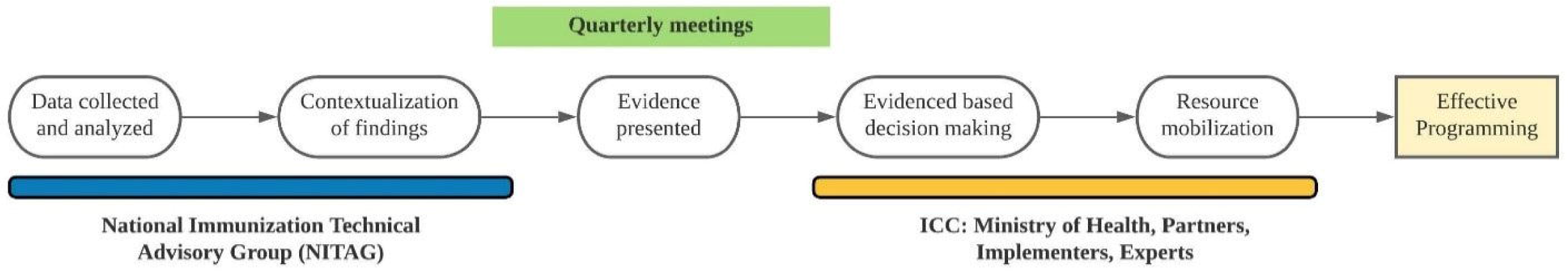
Programming accomplished through collaboration of the ICC and NITAG in Zambia.

**Table 3:** Comparing mandate and membership of ICCs and NITAGs.

|  | *ICC* | *NITAG* |
| --- | --- | --- |
| ***Mandate*** | Fosters collaboration between entities<br>Supports EPI resource mobilization<br>Supports advocacy efforts<br>Acts as a decision-making forum<br>Holds quarterly meetings<br>Lobbies for funding | Collects and analyses data to provide the MoH and ICC with evidence-based advice<br>Holds an advisory role<br>Develops and supports grant proposals<br>Provides technical advice<br>Supports contextualization |
| ***Membership*** | MoH (chair)<br>Representatives from technical groups<br>Donors and implementing partners<br>Civil Society Organizations<br>EPI officers (including sub-committee members and district reps),<br>Private sector | Core members include: experts in public health, health care, epidemiology, vaccine sector, sociology, economics<br>Liaison: Technical partners (WHO, UNICEF)<br>Secretariat: Ensures NITAG is functioning |

Through formal interactions, the ZITAG empowers the ICC to make evidence-based decisions. The ZITAG presents evidence to the ICC either recommending or discouraging a particular strategy or intervention based on context, formative research, piloting, and situational analysis. The MoH considers the evidence presented by the ZITAG when formulating national immunization policies and strategies. Integration of both bodies within national decision-making is enhanced by the distinct roles of each entity, a component that is necessary for the success of advisory and coordination committees [1,6]. Despite its recent establishment, Zambian officials interviewed for this study value the ZITAG’s ability to provide context-specific recommendations for immunization programming [14].

> “In terms of professional bodies in health, we have a good number of them that attend [regular coordination] meetings; also, we have the NITAG that has people pulled from different professions. So that’s quite a key component in the governance system.” (EPI personnel)

Additionally, the ZITAG includes several technical stakeholders, such as academics, epidemiologists, health care professionals, scientific societies, and technical experts from NGOs, whereas ICC membership includes a broader range of leadership.[14] Key informants in Zambia agreed that delays from coordination between the ICC and ZITAG are offset by the benefits of more robust, evidence-based decisions that are tailored to local context, which aligns with findings from existing literature.[7] In some cases, members of the ZITAG and EPI-TWG report questioning the efficiency and transparency of the ICC, which suggests that even further improvements in coordination and communication between these entities may be needed.

The addition of a fully functioning ZITAG, alongside the decision-making authority of the ICC, supports strategic planning and advocacy for funding:

> “The ZITAG is looking at [immunization] from a very critical standpoint, and from a very technical standpoint – looking at costs, disease burden, etc. I think [the ZITAG is] an external body that’s incredibly important to give recommendations compared to the ICC which [does not always include] many technical partners.” (CIDRZ personnel)

### 3.3 How does the Zambian ICC compare with the ICCs of other countries in Africa?

There is limited information available comparing the functionality of ICCs in different countries or assessing the interplay between ICCs and NITAGs. From our literature review, we found that Kenya and Tanzania, both with DTP3 coverage comparable to that of Zambia, have also experienced the benefits of expanding their ICC membership [18–22]. The Kenyan ICC also utilizes a multisectoral approach, which includes alignment of priorities with other health sectors, for crafting immunization policy through cooperation with the KENITAG. The ICC of Tanzania schedules additional meetings with its NITAG for strategizing efforts to reduce bottlenecks in delivery and improve uptake of immunization services [22]. While all countries receiving Gavi funding are advised to establish an ICC and NITAG, only some have reported fully utilizing collaboration of the two bodies for decision-making [19,23]. With active, inclusive, and collaborative ICCs and NITAGs, Zambia, Kenya, and Tanzania utilized these forums through defining clear roles and prioritizing the efficacy of immunization programs.

### 3.4 Transferable lessons

The structure and role of the Zambian ICC may act as a model for other countries by adapting based on the following transferable lessons:

- Expand ICC membership to include a variety of government officials, local representatives, community organizations, external partners, and donors to foster coordination and collaboration across stakeholders.
- Expand ICC mandate and scope to include other areas of health (i.e., reproductive, maternal, and child health) in order to foster integration of programs and health systems strengthening while still prioritizing immunization.
- Establish distinct and complementary roles for the ICC and NITAG to ensure efficient, collaborative, and sustainable integration. Continuously revisit the need for additional communication between groups.

## 4. Limitations

This study has several limitations. First, we focused on Zambia as a positive deviant in vaccine exemplars but were unable to carry out a similar analysis in a non-exemplary country to compare immunization coverage. Second, the research tools focused on the factors that drove catalytic change and did not probe on interventions or policies that were unsuccessful. Third, using qualitative methods to understand historical events was challenging; interviewees often spoke about current experiences rather than discussing historical factors. However, research assistants probed respondents to reflect on longitudinal changes in the immunization program.

## 5. Conclusion

Zambia’s status as an exemplar in vaccine delivery was likely supported by the strength, expansion, and prioritization of its national coordination and technical advisory committees. While most countries have functional ICCs, the Zambian ICC demonstrated enhanced strategic planning, decision-making, and lobbying efforts regarding vaccine programming. More research is needed to explore ICC functionality, compare ICCs from different countries, explore costs related to ICC activities, and assess the integral relationship between the ICC and NITAG. Findings from this paper may contribute to the decision-making processes, long-term engagement, membership, and mandates for ICCs in other countries.

## Conflict of interest statement

All authors declare: 1) No support from any organisation for the submitted work; 2) no financial relationships with any organisations that might have an interest in the submitted work in the previous three years; 3) no other relationships or activities that could appear to have influenced the submitted work.

## Contributors

KR, BD, WK, RB contributed to data collection; ZS, KR, KH, RD, BD, AE, KRI, SR, WO, WK, RB, MF contributed to analysis/interpretation; ZS, KR, KH, RD drafted the article; All authors critically revised the article. All authors provided approval of the submitted version. The views expressed are those of the authors alone and do not necessarily reflect the policies or recommendations of the institutions with whom they are affiliated.

## Declaration of interests

Authors declare no competing interests.

## Data Availability

Available upon request

## Acknowledgments

We thank the Center for Family Health Research in Zambia for their partnership in this study. In addition, we thank Sarah Chesemore, Anna Rapp, Tove Ryan, and Ethan Wong from the Bill and Melinda Gates Foundation; Kate Buellesbach, Nathaniel Gerthe, Gloria Ikilezi, Caitlyn Mason, David Phillips, and Oliver Rothschild from Gates Ventures; and the Vaccine Exemplars Research Advisory Group for their insights, specifically Agnes Binagwaho, Laura Craw, Carolina Danovaro, Anuradha Gupta, Heidi Larson, Penelope Masumbu, Kate O’Brien, Helen Rees, Lora Shimp, and Aaron Wallace. We gratefully acknowledge the participants who gave their time and insights to help us better understand Zambia’s vaccine delivery system.

## Funding statement

This work was supported by the Bill & Melinda Gates Foundation, grant number OPP1195041. Pilot and proposal development funds were provided by Gates Ventures.

## Data statement

The data are not publicly available as all data are confidential.

## Notes

### Competing Interest Statement

The authors have declared no competing interest.

### Funding Statement

Funding from the Bill and Melinda Gates Foundation.

### Author Declarations

The study was approved by the University of Zambia Biomedical Research Ethics Committee (Federal Assurance No. FWA00000338, REF. No. 166-2019), the National Health Research Authority in Zambia, and the Institutional Review Board committee of Emory University, Atlanta, Georgia, USA (IRB00111474).

### Summary of Updates

Changes to the structure of this manuscript were made due to feedback from journals following review.

